# Continuous monitoring of endotracheal tube obstructions using naturally occurring pressure and flow oscillations

**DOI:** 10.64898/2026.05.01.26352226

**Authors:** Ben Fabry, Christian Kuster, Roland Francis

## Abstract

The endotracheal tube resistance dominates the total airway resistance in most intubated patients. Mucus deposition and biofilm formation can rapidly increase tube resistance and thereby contribute to serious ventilatory impairments, including dynamic hyperinflation, intrinsic PEEP build-up, added work of breathing, and patient-ventilator asynchrony. During controlled mechanical ventilation, an increased tube resistance can be inferred from the difference between peak and plateau pressure, but this approach fails during pressure-supported spontaneous breathing. Here, we present a method that estimates the linear and nonlinear components of tube resistance from naturally occurring airway pressure and flow fluctuations at the airway opening, without a tracheal pressure sensor and without applying mandatory forced oscillations. This is achieved by solving the equation of motion using band-pass filtered airway pressure and flow signals. Band-pass filtering isolates the relevant resistive and inertive pressure losses across the tube by removing slow contributions from muscle pressure and lung elastance as well as high-frequency noise. The method accurately recovers both linear and nonlinear tube resistance parameters with < 10% error and < 2% bias. Moreover, it enables real-time implementation of full Automatic Tube Compensation (ATC), even in the presence of severe tube obstructions. Continuous estimation of endotracheal tube resistance from naturally occurring airway pressure and flow fluctuations enables real-time detection of clinically relevant tube narrowing and may help improve patient safety, reduce patient-ventilator asynchrony, and facilitate weaning.

## Introduction

In most intubated patients undergoing mechanical ventilation, including patients with acute respiratory distress syndrome (ARDS), the endotracheal tube (ETT) represents the major component of total airway resistance [1, 2]. The resistance of the lower airways (after the larynx) in healthy lungs is typically around 1 mbar/(L/s) [3]. By contrast, even under ideal conditions, a clean tube with inner diameter (I.D.) of 8 mm imposes a pressure drop of approximately 6.5 mbar at a flow of 1 L/s, and a clean 7 mm tube imposes a pressure drop of approximately 10 mbar at the same flow [2].

This added resistive load from the tube is a major source of added work-of-breathing during pressure-supported modes of spontaneous breathing [4]. It also limits expiratory flow, increases intrinsic PEEP, and contributes to trigger failure and patient-ventilator desynchronization [5]. Clinical studies have shown that the resistance of a clean tube, which is substantial even under normal unobstructed conditions, often increases markedly after intubation, sometimes within the first 30 minutes [1, 6]. Reasons for this increased resistance include kinking, mucus accumulation, and biofilm formation. On average, the tube resistance rises by about 69% after 3 days [1], and even a doubling or tripling of the tube resistance above baseline has been reported [1, 6]. Such severe tube obstruction can substantially worsen tube-related ventilatory impairments and may lead to weaning failure.

During volume-controlled ventilation, total airway resistance can be inferred from the difference between peak and plateau airway pressure, and severe tube obstructions are readily detectable. By contrast, no such estimate is available during pressure-supported spontaneous breathing. Progressive tube obstructions may therefore go unnoticed.

In principle, total airway resistance during pressure-controlled ventilatory modes could be assessed using forced oscillometry, which superimposes high-frequency flow oscillations and measures the resulting pressure response [7]. However, this approach requires dedicated hardware and assumes a linear pressure-flow relationship for resistive losses, which does not apply to endotracheal tubes. The tube resistance is strongly flow-dependent due to the mostly turbulent flow conditions inside the tube, and due to flow separation at the transition into the trachea and the high curvature at the swivel connector [8].

In this study, we present a method for real-time estimation of the linear and nonlinear components of endotracheal tube resistance from airway pressure and flow measurements. The method does not require forced oscillations or a tracheal pressure sensor. The central idea is that naturally occurring mid-frequency (order of 10 Hz) flow fluctuations generated by the ventilator produce corresponding airway pressure fluctuations due to the resistive and inertive properties mainly of the endotracheal tube. Using band-pass filtering, these mid-frequency pressure fluctuations can be separated from slower pressure fluctuations caused by spontaneous breathing effort and elastic recoil of the respiratory system, as well as from faster pressure fluctuations due to sensor noise, acoustic phenomena, and higher-order effects. We validated the method in a test-lung setup with adjustable tube narrowing, compared the estimated parameters with independently measured reference values, and show that the method can track evolving tube obstruction, which is a prerequisite for automatic compensation of tube resistance.

## Methods

### Experimental setup

Original-length endotracheal tubes (Rüsch Super Safety Clear, Teleflex, Ireland; inner diameter 5, 7, 7.5, 8, 8.5 or 9 mm) were inserted and cuffed airtight into an artificial trachea made of Plexiglas with an inner diameter of 2.1 cm (Fig. 1). Tracheal pressure was measured through 12 small radial holes that were equally spaced around the circumference of the artificial trachea. The measurement site was located 60 mm distal to the tube tip and 50 mm proximal to the distal end of the artificial trachea. Airway pressure was measured through 8 radial holes in a short tube between the swivel connector and the flow sensor.

**Fig. 1.**
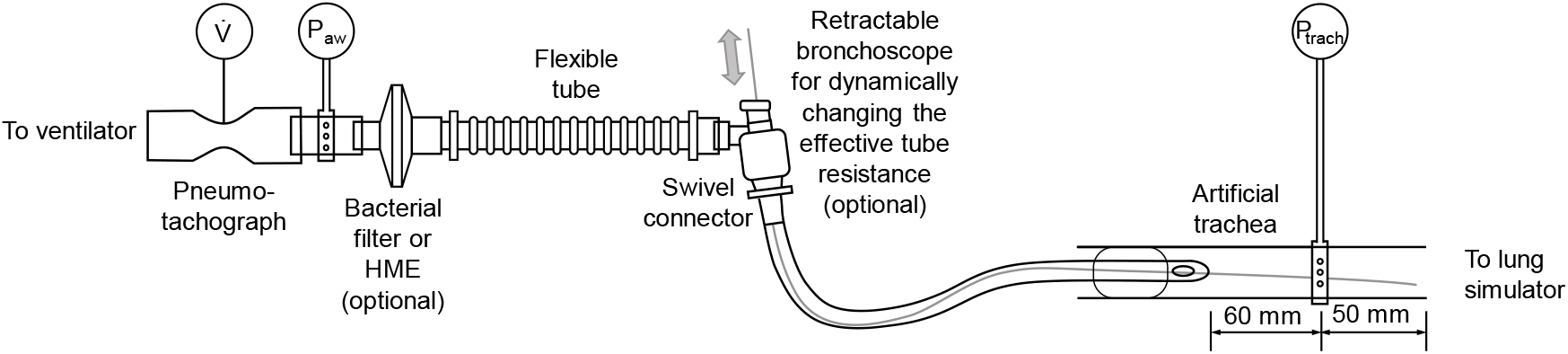
Experimental setup for measuring tube resistance. The endotracheal tube was inserted into an artificial trachea. To simulate an obstruction, a bronchoscope was advanced through the tube beyond the tracheal pressure measurement site. Tracheal pressure was measured only to validate the resistance values estimated from airway pressure and flow fluctuations.

Airway and tracheal pressures were measured with piezo-resistive differential pressure sensors (Honeywell HCS series, USA; range ±80 mbar). Gas flow was measured with a factory-calibrated thermal mass flow sensor (SFM3300, Sensirion, Switzerland). Pressure and flow signals were sampled at 250 Hz. In selected measurements, a bacterial filter or heat and moisture exchanger was placed between the flow sensor and the tube; in those cases, the measured resistive segment included that component as well as the tube.

To simulate progressive obstruction, a disposable bronchoscope (aScope 4 Broncho, Ambu, Denmark; shaft diameter 5 mm) was introduced through the swivel connector and advanced into the tube. The bronchoscope shaft extended beyond the tracheal pressure measurement site so that the reference pressure drop across the measured segment remained well defined.

The artificial trachea was connected to a custom-built lung simulator that consisted of a 20 L glass container and a 2.8 L piston pump, as described previously [9]. A stepper-motor drive controlled the motion of a piston so that the simulator could reproduce a prescribed airway resistance, respiratory-system elastance, and time-varying muscle pressure. Unless stated otherwise, the simulator breathed spontaneously with inspiratory time 1 s, expiratory time 2 s, maximum inspiratory muscle pressure 10 mbar, airway resistance 2 mbar/(L/s), and respiratory-system elastance 20 mbar/L (compliance 50 ml/mbar).

The setup was connected either to a commercial intensive care ventilator (Evita V600, Dräger, Germany) or to a prototype ventilator based on a system described previously [10]. Pressure support was varied between 0 and 20 mbar above PEEP 5 mbar or 0 mbar. Automatic Tube Compensation was evaluated with settings that matched the estimated tube resistance parameters.

### Signal processing and model formulation

We start from the classical equation of motion of the respiratory system, 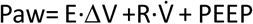, where E is the total elastance and R is the total resistance of the respiratory system, 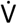 is the gas flow, PEEP is the end-expiratory (airway) pressure, and ΔV is the change in lung volume above the end-expiratory lung volume at the given PEEP. In intubated patients, R becomes flow-dependent according to 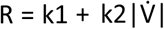, where k1 and k2 are the linear and nonlinear Rohrer coefficients, respectively [3]. Flow acceleration within the endotracheal tube can also cause appreciable inertial pressure losses, described by 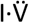, with inertance I and flow acceleration 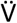. If the patient breathes spontaneously, the airway pressure includes contributions from the typically negative pressure generated by the respiratory muscles, Pmus. Taken together, the equation of motion becomes

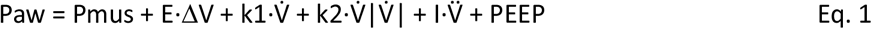

Airway pressure Paw and gas flow 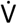 were sampled at 250 Hz. Flow acceleration 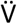 was obtained by numerical differentiation, and volume V was obtained by numerical integration of the raw flow signal.

In the following, we describe three approaches to estimate the resistance and inertance terms:

### Time difference analysis

During pressure-supported spontaneous breathing, the muscle pressure Pmus fluctuates but is unknown. However, over a short time interval corresponding to the sampling period dt (4 ms), the muscle pressure can be assumed to remain approximately constant. Therefore, its contribution cancels when taking the difference between consecutive samples. This yields a time-difference form of the equation of motion:

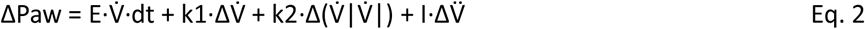

Here, Δ denotes the difference between signals sampled 4 ms apart. Before taking the difference, all signals were digitally low-pass filtered using a zero-phase 4th-order Bessel filter with a cutoff frequency of 25 Hz to suppress high-frequency sensor noise as well as acoustic and higher-order effects while preserving physiologically relevant fluctuations. For each pair of consecutive samples, Eq. 2 yields one linear equation. Stacking these equations over one or more breaths results in an overdetermined system, which was solved by least-squares linear regression to estimate E, k1, k2, and I. Because the change in elastic recoil pressure 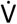·dt·E between two sampling time points is small, the elastance E during spontaneous breathing cannot be reliably estimated. Hence, the term E·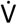·dt can be omitted.

### Band-pass filtering

Taking the time difference of low-pass-filtered signals yields a band-pass-like response. An alternative approach is therefore to insert band-pass-filtered measured signals into Eq. 1, stack the resulting equations from one or more breaths, and solve the overdetermined system by least-squares linear regression to estimate E, k1, k2, and I. Unless noted otherwise, we used a zero-phase 4th-order Butterworth band-pass filter with a low cutoff frequency of 3.7 Hz to remove slow contributions from the unknown muscle pressure and elastic recoil pressure fluctuations, and a high cutoff frequency of 40 Hz to remove fast contributions from sensor noise, acoustic, and higher-order effects.

### Bootstrapping

Bootstrapping employs the same principle of separating the relevant resistive and inertive mid-frequency pressure fluctuations from low-frequency muscle pressure fluctuations and high-frequency noise. The bootstrapping method iteratively optimizes E, k1, k2, and I to minimize the least-squares residuals between measured and fitted airway pressure, using bounded nonlinear least-squares optimization with a trust-region reflective algorithm. Starting from initial values of E, I, k1, and k2, we first estimated Pmus by rearranging Eq. 1 and inserting the unfiltered measured signals. This estimate of Pmus was then low-pass filtered with a zero-phase 4th-order Butterworth filter with a cutoff frequency of 3.2 Hz and inserted back into Eq. 1, hence the name bootstrapping. The resulting airway pressure estimate was low-pass filtered at 40 Hz and compared with the equally filtered measured airway pressure to compute the residuals. The updated parameters were then used as starting values for the next iteration until convergence.

Accuracy was assessed by comparing the directly measured pressure drop across the endotracheal tube,

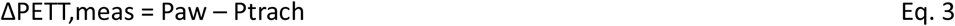

with the pressure drop calculated from the estimated tube parameters,

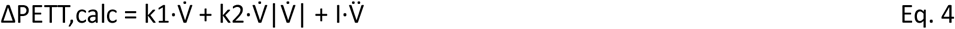

## Results

Fig. 2 shows representative examples from situations in which a lung simulator was connected to a ventilator via endotracheal tubes of different sizes, with or without a bacterial filter. In all cases, the method estimated the linear and nonlinear components of endotracheal tube resistance with high accuracy, as verified against directly measured reference values. Accordingly, the predicted tracheal pressure and pressure-flow loop agreed closely with the measured values.

**Fig. 2.**
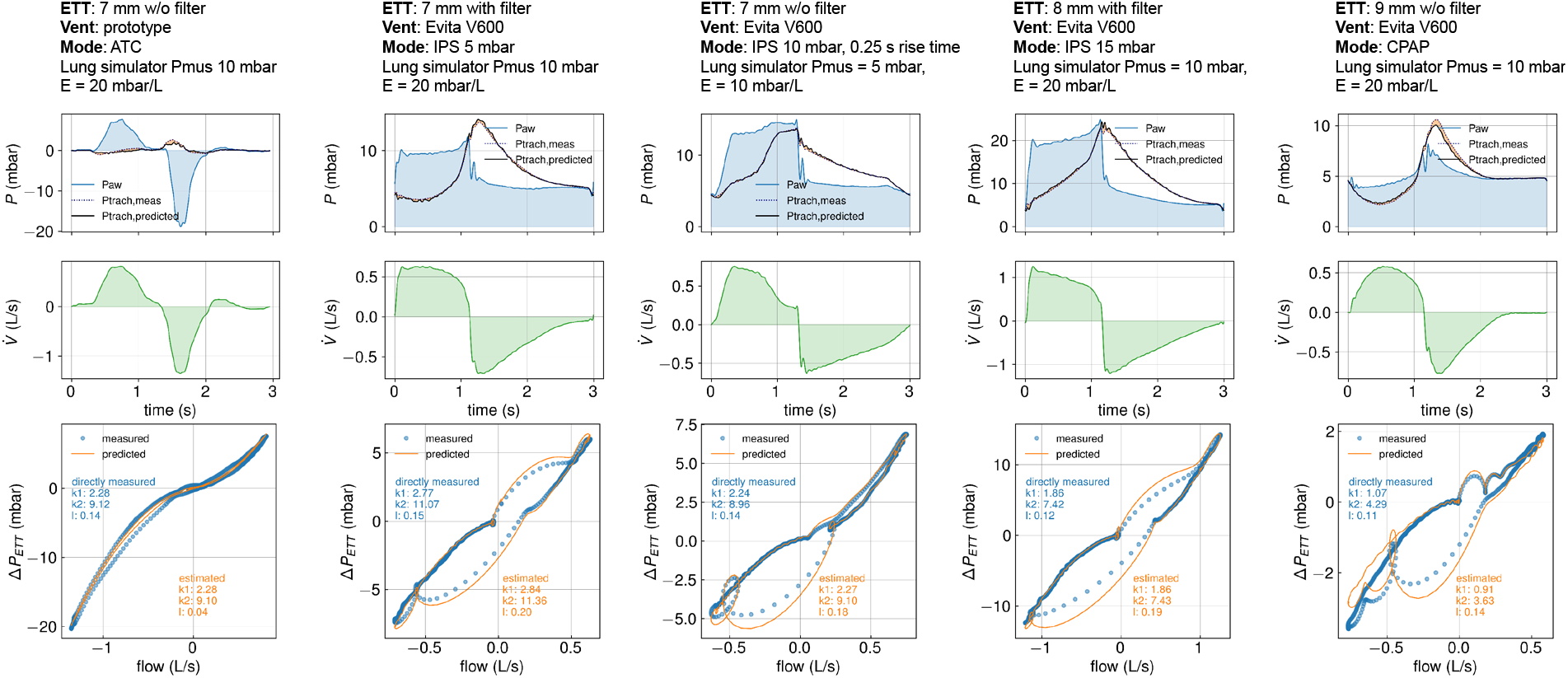
Performance of the method under different conditions. Each panel shows measured airway pressure, measured and estimated tracheal pressure (top), flow (middle), and measured and estimated pressure drop across the endotracheal tube, including the bacterial filter if present (bottom), during a single breath. The predicted pressure drop, ΔPETT,calc, was calculated using Eq. 4 from tube resistance and inertance parameters estimated by the band-pass filtering method from that breath. Estimated tracheal pressure was then obtained as Ptrach,predicted = Paw - ΔPETT,calc. Flow was delivered either by a commercial ventilator (Evita V600) or by a prototype ATC ventilator via endotracheal tubes of different sizes, with or without a bacterial filter, as indicated in the figure.

During patient-triggered pressure support, pressure and flow fluctuations of sufficient magnitude to estimate tube resistance occurred predominantly at the beginning and end of the pressure-support cycle. As the level of pressure support approached zero, the magnitude of these fluctuations decreased and the accuracy of tube resistance estimation declined (Fig. 3). The time-difference method in particular tended to underestimate tube resistance at low pressure-support levels. The band-pass and bootstrapping methods performed well even at zero pressure support (CPAP) for tube diameters of 8 mm or less. For larger tube diameters, both methods also underestimated the tube resistance at pressure-support levels below 5 mbar.

**Fig. 3.**
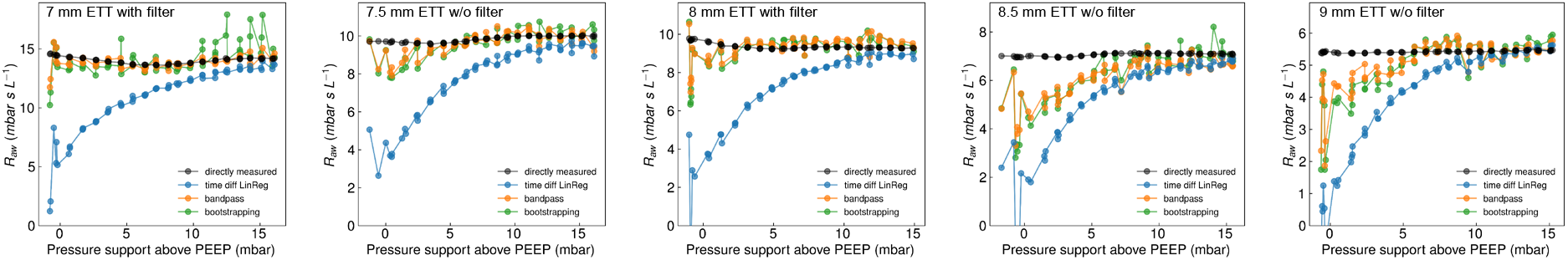
Performance of the method at different pressure-support levels. Directly measured (black) and estimated (colored) tube resistance values at a flow of 1 L/s are shown as a function of pressure-support level during patient-triggered inspiratory pressure support, for endotracheal tubes of different diameters with or without a bacterial filter. Tube resistance was estimated with three different methods. With increasing pressure support, the estimated values approached the directly measured resistance. Each dot represents data from one breath. The breathing pattern was generated by a lung simulator with standard settings as described in Methods.

To test whether the method can detect a gradual increase in tube resistance, we inserted a 5 mm bronchoscope stepwise into a 7 mm endotracheal tube during spontaneous breathing of the lung simulator, with maximum inspiratory muscle pressure set to 5 mbar and inspiratory pressure support set to 10 mbar above PEEP. The estimated tube resistance closely tracked the directly measured resistance over the full range, including extreme values when the bronchoscope was fully advanced beyond the tip of the tube (Fig. 4).

**Fig. 4.**
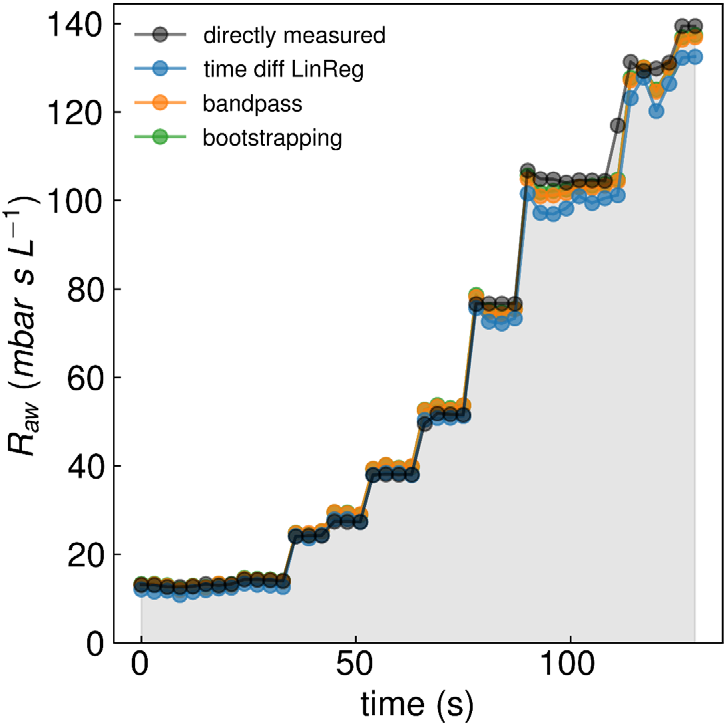
Continuous detection of tube obstruction. Measured and estimated tube resistance at a flow of 1 L/s during progressive obstruction of a 7 mm endotracheal tube by a 5 mm bronchoscope. Jumps in the data indicate time points at which the bronchoscope was advanced rapidly by several centimeters. Each symbol represents data from one breath.

We next tested the method on a larger dataset comprising 500 breaths measured under a variety of ventilation modes, ventilator settings, and experimental conditions. These included continuous positive airway pressure (CPAP) with or without automatic tube compensation (ATC), patient-triggered inspiratory pressure support ventilation, pressure-support levels of 0–20 mbar, pressure rise times of 0 or 250 ms, lung simulator efforts of Pmus = 5 or 10 mbar, endotracheal tube diameters of 5.0, 7.0, 7.5, 8.0, 8.5, and 9.0 mm, and measurements with or without a bacterial filter. Across these conditions, the band-pass and bootstrapping methods showed mean absolute percentage errors below 10%, median absolute percentage errors below 5%, and mean bias below 2% relative to directly measured tube resistance values. By contrast, the time-difference method showed larger errors and a marked negative bias, because it underestimated resistance when pressure support and fluctuation amplitude were small (Table 1).

**Table 1.** Comparison of the three analysis variants across the full dataset, expressed as mean absolute percentage error, median absolute percentage error, and mean bias relative to directly measured tube resistance values.

|  | time difference | band-pass | bootstrapping |
| --- | --- | --- | --- |
| Mean absolute error (%) | 20.2 | 6.9 | 7.0 |
| Median absolute error (%) | 9.8 | 3.8 | 4.1 |
| Mean bias (%) | -9.7 | -1.2 | -1.1 |

## Discussion

This study shows that flow fluctuations generated by the demand-flow valve during spontaneous breathing are sufficient to estimate endotracheal tube resistance in situ and to track its changes over time. The method is based on the principle that airway pressure fluctuations in the frequency range of about 4-40 Hz are dominated by the resistance and inertance of the endotracheal tube. Band-pass filtering removes slower pressure variations arising from spontaneous breathing effort and respiratory-system recoil, as well as higher-frequency noise. Because the low-frequency components are removed, respiratory-system elastance could not be estimated reliably. In addition, the estimated inertance typically differed from the directly measured inertance by more than 10%. However, this does not compromise the ability of the method to estimate tube resistance and its changes over time.

We validated the method in a laboratory test-lung setup comprising a rigid-wall artificial trachea, adjustable flow resistance and lung elastance, and different breathing patterns. An increase in tube resistance was simulated by introducing a bronchoscope into the tube lumen. In this setup, the method estimated tube resistance with high accuracy, with mean deviations of less than 10% from directly measured reference values. Because the method can estimate tube resistance from a single breath, it can detect sudden tube obstruction and track ongoing changes in tube resistance in real time (Fig. 4).

A limitation of this study is that the method was not tested in ventilated patients, because the prototype ventilator and the flow-pressure measurement unit were not clinically approved. We therefore cannot verify that the method performs as intended under certain clinical conditions, for example when tube resistance is increased by a mucus plug that changes position during the breathing cycle, or when the patient’s airways partially close during expiration.

A further limitation is that the method yields accurate values only when the flow fluctuations generated by the demand-flow valve are of sufficient amplitude in the frequency range of 4-40 Hz. When the demand-flow valve is located near the patient, as in the prototype ATC ventilator, this condition was usually met. However, when the patient is connected to a conventional ICU ventilator, the long tubing can dampen these fluctuations, which may then become too small for the method to perform reliably. For example, during patient-triggered inspiratory pressure support, a support level of at least 5 mbar above PEEP was required to generate sufficient flow and pressure fluctuations for accurate resistance estimates in tube diameters above 8 mm (Fig. 3). In principle, however, commercial ventilators could superimpose small forced oscillations to assess ETT resistance intermittently when naturally occurring fluctuations from the demand-flow valve are insufficient.

In summary, the method described here enables continuous in situ monitoring of endotracheal tube resistance and reliable detection of tube obstruction, including additional resistance contributions from bacterial or HME filters, or from inserted bronchoscopes. It offers a practical approach to enhance patient safety and improve the management of ventilated patients.

## Declarations

### Author approval

All authors have seen and approved the manuscript.

### Ethics approval and consent to participate

This study did not involve human participants, data, or identifiable images.

### Consent for publication

Not applicable. This study did not involve human participants, data, or identifiable images.

### Availability of data and materials

The raw data and analysis software can be obtained from the corresponding author upon request.

### Competing interests

BF and CK are the inventors of a pending patent application (PCT/EP2023/074318; WO2024052339A1) related to the prototype ventilator described. BF is the inventor of a pending patent application (EP26160568.7) related to the method described. RF declares no competing interests.

### Funding

This study was not funded by specific project grants.

### Author Contributions

BF designed the study, CK designed and built the active lung simulator, BF and CK designed and built the prototype ventilator, BF performed the experiments and analyzed the data, BF and RF wrote the manuscript.

